# Recall-by-Genotype of Neurodevelopmental Disorder Copy Number Variants from a Multi-Ancestry, Healthcare-System Biobank

**DOI:** 10.1101/2024.07.16.24310489

**Authors:** Nina Zaks, Behrang Mahjani, Abraham Reichenberg, Rebecca Birnbaum

## Abstract

**Background:** Clinical biobanks linking electronic health records (EHRs) with genotype data are expanding, enabling investigation of genomic risk factors for psychiatric disorders. However, few recall-by-genotype (RbG) studies have been published—particularly for psychiatric risk variants in diverse healthcare systems—indicating a need for further research to inform implementation. Some rare copy number variants (CNVs) confer substantially increased risk for neurodevelopmental disorders (NDDs) and cognitive impairment. We recalled NDD CNV carriers from Bio*Me*, a multi-ancestry biobank within the Mount Sinai Health System, for in-depth phenotyping and empirical insights into the implementation of RbG in psychiatry.

**Methods:** From BioMe, 892 adults were recontacted: 335 NDD CNV carriers, 217 with schizophrenia, and 340 neurotypical controls. Of these, 18% responded to recontact, 12% were screened for participation, and 10% began the study. Participants completed structured clinical and cognitive assessments.

**Results:** Seventy-three participants (8% of those recontacted) completed the study: 30 NDD CNV carriers, 20 schizophrenia cases, and 23 controls. The mean age was 48.8 years, 66% were female, and ancestry was 37% African, 34% Hispanic, and 26% European. Seventy percent of NDD CNV carriers had at least one neuropsychiatric or developmental condition, including 40% with mood or anxiety disorders. Among 22 NDD CNV carriers at loci previously examined for cognitive effects, performance was impaired on digit span backward (*β*= –1.76, FDR = 0.04) and sequencing (*β*= –2.01, FDR = 0.04) compared with controls but outperformed schizophrenia cases on verbal learning (*β*= 4.5, FDR = 0.05).

**Conclusions:** This proof-of-concept RbG study of rare psychiatric risk variants from a multi-ancestry biobank demonstrates both opportunities and challenges for recontact within healthcare systems. Despite modest enrollment, recalling individuals—including those affected by psychiatric illness and cognitive impairment—yielded a genotypically defined cohort and phenotypes not captured in EHRs, underscoring the potential of RbG to advance precision psychiatry.

## INTRODUCTION

Clinical biobanks, derived from healthcare systems or population samples, that link electronic health records (EHRs) to genotype data are rapidly expanding, offering opportunities to investigate disease-relevant genomic risk factors.^1^ Compared with hundreds of published *in silico* analyses and computational approaches leveraging biobank data, relatively few recall-by-genotype (RbG) studies —requiring active recontact of participants —have been published, and their feasibility and implementation remain uncertain.^2, 3^ Biobank-based RbG provides a potentially powerful approach to recruit individuals carrying disease-relevant genetic variants for observational or interventional studies.

For example, some rare copy number variants (CNVs)—genomic microdeletions or microduplications greater than 1kb in length —confer markedly increased risk for multiple neurodevelopmental disorders (NDDs) and are among the highest-effect genetic risk factors in neuropsychiatry.^4–9^ Collectively, NDD CNVs account for up to 20% of autism spectrum disorder cases, 14% of developmental delay and intellectual disability, and 1–3% of schizophrenia cases, contributing an estimated 1–3% of heritability across psychiatric disorders.^9–14^ NDD CNVs also negatively affect neurocognition, a quantitative trait linked to functional outcomes, as reported in clinically ascertained cohorts.^10, 15^ Prior studies in population samples—including the Icelandic population cohort and UK biobank—demonstrated cognitive effects even among neurotypical individuals without psychiatric illness. ^15–19^ NDD CNVs exhibit variable penetrance and expressivity, resulting in heterogeneous outcomes ranging from unaffected to severely affected.^20–22^ Although individual NDD CNVs are rare, their combined prevalence may reach up to 2% of population cohorts.^18, 19, 23–25^ Identifying NDD CNV carriers could be valuable for elucidating the phenotypes of disease-relevant variants and for informing targeted interventions or disease management.

The feasibility of recalling rare CNV carriers from biobanks remains uncertain. Factors influencing success may include willingness to participate in research, logistical considerations, engagement with the healthcare system, sociodemographic and cultural factors, or perceived study relevance. A previous study within the Geisinger *MyCode* Community Health Initiative identified carriers of 31 neuropsychiatric CNVs and reported positive reactions to genetic disclosure for 141 carriers of nine CNVs. However, the recontacted cohort was predominantly European, derived from a relatively contained healthcare system, and did not include cognitive analyses.^24^ A recent analysis of the Bio*Me* biobank, a multi-ancestry biobank derived from the Mount Sinai healthcare system, reported a 2.5% prevalence of NDD CNVs among approximately 25,000 adults with a mean age of 50 years.^26^ Enrichments were observed for congenital disorders and major depressive disorder, as well as for several medical conditions, including hypertension, obesity, and increased body mass index. That report, however, was limited to EHR outcomes. To address this gap, we conducted a proof-of-concept recall-by-genotype study of rare NDD CNV carriers and comparators idiopathic schizophrenia and neurotypical controls without NDD CNVs.. This study aimed to characterize clinical and cognitive phenotypes beyond EHR data and to provide empirical observations relevant to implementing RbG for psychiatric risk variants.

## METHODS

### IRB Approval

The Bio*Me* biobank is an EHR-linked resource of genotyped individuals of diverse ancestry, recruited across the Mount Sinai Health System (New York, NY) since 2007 and approved by the Institutional Review Board (IRB) at the Icahn School of Medicine at Mount Sinai. ^26, 27^ As previously described, biobank participants were recruited across the healthcare system, mostly from various outpatient medicine clinics, with some recruitment from outpatient psychiatry clinics .^26^ For the present study, the IRB approved Bio*Me* data access, participant recontact, and study assessments, without return of genetic results. The study was conducted from November 12, 2020, to March 23, 2023.

### Recruitment

Within Bio*Me*, we identified three groups of adults, ages 18-65, for recontact. The age range was selected to focus on adults while minimizing potential confounding from age-related cognitive decline: (i) **NDD-CNV carriers**, harboring at least one NDD CNV^26^; (ii) **Schizophrenia**, defined by at least two International Classification of Diseases (ICD10) codes for schizophrenia and without NDD CNVs; (iii) **Neurotypical controls**, without ICD-10 code for neuropsychiatric disorders and without NDD CNVs. Respondents were excluded for unstable or severe medical illness, substance or alcohol use disorder, or conditions affecting cognition. Chart review was conducted by a board-certified psychiatrist (RB). (Supplementary Methods). Consistent with the prior Bio*Me* NDD CNV report, ancestry was determined by self-report. Participants who had previously consented to be recontacted were contacted for the current study via mail, email, or phone. All eligible NDD CNV carriers and all eligible individuals with schizophrenia were invited, along with a subset of matched neurotypical controls.

In total, 892 participants from Bio*Me* meeting eligibility criteria were recontacted: 335 NDD CNV carriers, 217 with schizophrenia, and 340 controls (Figure 1). The initial response rate was 18% (15% of NDD CNV carriers, 20% of schizophrenia cases, 19% of controls) 12% were screened for the study, and 10% started the study. The attrition from initial response to study participation was due to unreachable participants or lack of interest (6%), reported alcohol or substance use (1%), or medical illness confounding cognition (1%). An additional 2% of enrolled participants began the study but were excluded because schizophrenia or control status could not be confirmed due to discordance between EHR and study assessment. No significant differences were observed between the comparator groups at any stage of recruitment: Responded (χ² = 2.17, p = 0.34), Screened (χ² = 2.97, p = 0.23), Started Assessments (χ² = 0.84, p = 0.66), or Completed Assessments (χ² = 1.49, p = 0.48).

**Figure 1.**
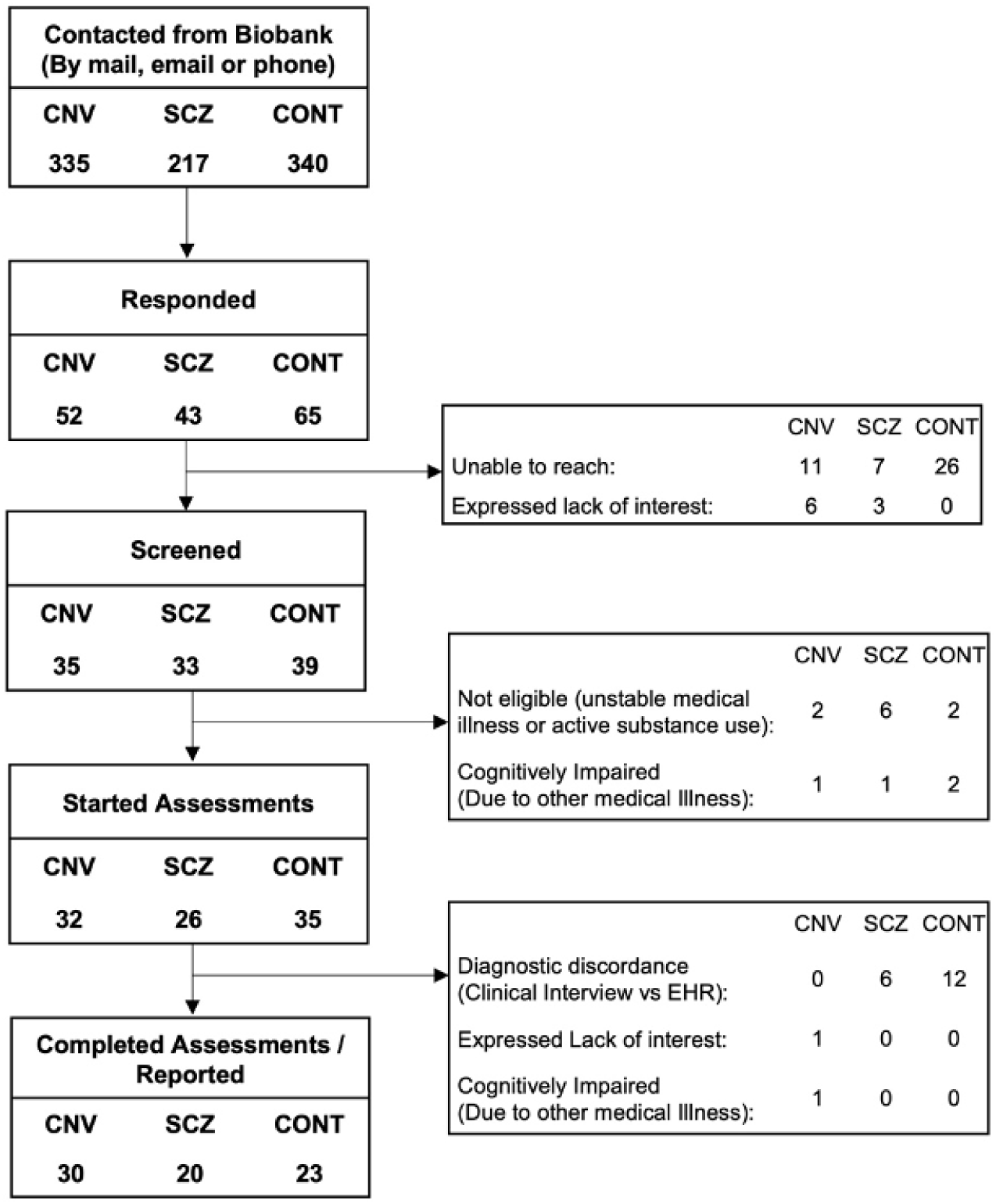
Study Recruitment. Flow diagram showing recontact of 892 Bio*Me* biobank participants across three comparator groups—neurodevelopmental CNV carriers, schizophrenia, and controls—from initial contact to completion of study assessments.

### Clinical and Cognitive Assessments

Assessments were performed remotely (Supplementary Figure 1, Supplementary Methods). The 30-minute clinical evaluation included the Mini International Neuropsychiatric Interview (MINI v7.0.2) for DSM-5 diagnoses, along with a medical, psychiatric, and developmental history, administered by a board-certified psychiatrist (RB).^28^ Cognitive assessments were administered by trained staff (NZ) using standardized instructions and scoring for comparability with published studies. A 30-minute neurocognitive battery included seven subtests: Digit Span (Forward, Backward, Sequencing) for attention and executive function; Category Fluency for processing speed and semantic memory; the Mayer-Salovey-Caruso Emotional Intelligence Test (Managing Emotions branch) for social cognition; and the Hopkins Verbal Learning Test–Revised (HVLT-R) immediate and delayed recall for verbal learning and memory (Supplementary Figure 1).^29, 30^ Each subtest targeted cognitive domains relevant to NDD CNVs and has been validated in both neuropsychiatric and neurotypical populations. Evidence supports the validity of remote administration in schizophrenia and other clinical and non-clinical populations (Supplementary Methods). ^31, 32^

### Statistical Analyses

Cognitive raw scores were modeled using linear regression to compare groups, with age, sex, and ancestry included as covariates. All analyses were performed in R 4.0.4. Statistical significance was evaluated using Benjamini-Hochberg false discovery rate (FDR) method.

## RESULTS

### Study Sample, Demographics

(Table 1, Supplementary Table 2): Overall, 73 participants (8% of those recontacted) completed the study: 30 NDD CNV carriers, 20 individuals with schizophrenia, and 23 controls. The mean age was 48.8 ± 10.2 years, 66% were female, and participants were of diverse self-identified ancestry, 37% African, 34% Hispanic, and 26% European. Age and ancestry did not differ significantly between groups. NDD CNV carriers were predominantly female (87%), whereas the schizophrenia group was mostly male (55%) (x^2^=5.9, p = 0.003). For the NDD CNV group female sex was associated with higher participation (x^2^=5.9, p=0.02), and for individuals with schizophrenia, older age was associated with higher participation (t-statistic=2.03, p=0.05) (Supplementary Table 1)

**Table 1.** Study Sample. (A) Demographic characteristics of the three study groups who were recontacted and completed assessments. (B) Copy-number-variant (CNV) loci of the 30 NDD CNV carriers who completed assessments.

|  | Biobank Participants (n=24,877) | NDD CNV (n=627) | NDD CNV (n=335) | Schizophrenia (n=217) | Controls (n=340) | CNV (n=30) | Schizophrenia (n=20) | Controls (n=23) | Test statistic | P-value |
| --- | --- | --- | --- | --- | --- | --- | --- | --- | --- | --- |
| Mean Age, Biobank Enrollment (SD) | 50.5 (17.3) | 50.4 (17.0) | 39.0 (10.8) | 45.4 (9.9) | 41.7 (10.7) | 37.3 [9.2] | 41 [9.3] | 39.9 [9.4] |  |  |
| Mean Age at Study Assessment (SD) |  |  |  |  |  | 47.3 [9.5] | 50.2 [10.1] | 49.5 [11.2] | 0.55 | 0.58 |
| Female (%) | 14,586 (59%) | 389 (62%) | 219 (65%) | 97 (45%) | 209 (61%) | 26 (87%) | 8 (40%) | 14 (61%) | 11.96 | 0.003 |
| Ancestry |  |  |  |  |  | - | - | - | 4.98 | 0.55 |
| African | 5965 | 168 | 95 | 101 | 111 | 10 (33%) | 9 (45%) | 8 (35%) | 0.77 | 0.68 |
| European | 7892 | 208 | 102 | 17 | 52 | 10 (33%) | 2 (10%) | 7 (30%) | 3.73 | 0.16 |
| Hispanic | 8536 | 200 | 109 | 87 | 151 | 9 (30%) | 9 (45%) | 7 (30%) | 1.42 | 0.49 |
| Other | 2484 | 51 | 32 | 12 | 26 | 1 (3%) | 0 (0) | 1 (4%) | - | - |

(B) NDD CNV Carriers, by CNV Locus
| NDD CNV | hg19 coordinates | Completed Study (n=30) |
| --- | --- | --- |
| TAR_dup | chr1:145,394,955-145,807,817 | 3 |
| 1q21.1del** | chr1:146,527,987-147,394,444 | 1 |
| 1q21.1dup | chr1:146,527,987-147,394,444 | 1 |
| 2q13del(NPHP1) | chr2:110,862,716-110,983,948 | 2 |
| 2q13dup(NPHP1) | chr2:110,862,716-110,983,948 | 3 |
| 15q11.2del | chr15:22,805,313-23,094,530 | 5 |
| 15q11.2dup | chr15:22,805,313-23,094,530 | 4 |
| 15q13.3dup(CHRNA7) | chr15:32,017,070-32,453,068 | 2 |
| 16p13.11del | chr16:15,511,655-16,293,689 | 2 |
| 16p13.11dup | chr16:15,511,655-16,293,689 | 2 |
| 16p11.2distal_del** | chr16:28,823,196-29,046,783 | 1 |
| 16p11.2del | chr16:29,650,840-30,200,773 | 1 |
| 17p12del | chr17:14,141,387-15,426,961 | 1 |
| 22q11.2del** | chr22:19,037,332-21,466,726 | 1 |
| 22q11.2dup | chr22:19,037,332-21,466,726 | 1 |

Notes: *p* values for age, sex, and ancestry reflect comparisons across the three groups. An *F* statistic was used for mean age; chi-square tests for sex and ancestry. Ancestry was self-reported. Thirty NDD CNV carriers completed the study; cognitive analyses were restricted to 22 carriers to match previously reported UK Biobank analyses, excluding four loci (15q13.3 [CHRNA7] duplication, 17p12 deletion, and 2q13 [NPHP1] deletion/duplication).18,19 \*\* = NDD CNV loci previously reported as significantly associated with schizophrenia.5 See Supplementary Table 2 for additional details on recontacted versus completed CNV carriers.

### Study Sample, Healthcare System Utilization

Overall healthcare utilization did not affect recruitment: the study sample averaged 125 clinical encounters versus 133 for other recontacted participants (t = 0.30, p = 0.76). However, recent engagement did influence participation, as study participants had, on average, a clinical encounter 12 weeks prior to recontact compared to 57 weeks for non-participants (t = 7.1, p = 1.1 × 10⁻¹⁰).

### Study Sample, NDD CNV Status

Of 30 NDD CNV carriers, 16 harbored duplications and 14 deletions, across 15 distinct NDD CNV loci; no individuals harbored multiple NDD CNVs (Table 1, Supplementary Table 2). Some NDD CNV loci were enriched among participants who completed assessments (i.e. TAR duplication, 15q11.2 deletion and 16p13.11 deletion) (Supplementary Table 2). None of the participants with schizophrenia nor neurotypical controls harbored NDD CNVs.

### Clinical Assessment, Results

MINI administration confirmed schizophrenia diagnoses and the absence of psychiatric illness in neurotypical controls (Figure 1). Of the 30 NDD CNV carriers, no cases of schizophrenia were identified, but one participant had bipolar disorder with psychosis. Mood or anxiety disorders were diagnosed in 40% of NDD CNV carriers, including five with major depressive disorder and two with obsessive-compulsive disorder. Clinical assessment findings were often discordant with or under-reported in EHRs, particularly for developmental history, much of which was not captured in the EHR (Table 2). For example, childhood speech delay warranting speech therapy was reported in three NDD CNV carriers, and one NDD CNV carrier reported global developmental delay including motor skills of walking. Further, formally diagnosed learning disorders were reported by approximately 30% of NDD CNV carriers, and a history of special education was reported for 13%, from elementary school through high school, across multiple years and multiple subject areas.

**Table 2.** Prevalence of Selected Clinical Features. Neuropsychiatric and neurodevelopmental features identified through structured assessments across the three study groups.

| CLINICAL FEATURE | NDD CNV (n=30) |  |  | SCZ (n=20) |  | CONT (n=23) |  |
| --- | --- | --- | --- | --- | --- | --- | --- |
|  | N | NDD CNV LOCI | EHR Unreported/<br>Discordant | N | EHR Unreported/<br>Discordant | N | EHR Unreported/<br>Discordant |
| MOOD OR ANXIETY DISORDER | 12 | TAR dup, 1q21.1 del, 1q21.1 dup, 2q13 (NPHP1) dup, 2q13 (NPHP1) dup, 15q11.2 dup, 15q11.2 dup, 15q13.3 dup (CHRNA7), 16p11.2 distal del, 16p13.11 del, 16p13.11 del, 16p13.11 dup | 6 | 0 | 0 | 0 | 0 |
| SEIZURE DISORDER | 1 | 16p11.2 distal del | 1 | 1 | 1 | 0 | 0 |
| DEVELOPMENTAL DELAY (SPEECH OR MOTOR) | 4 | 1q21.1 del, 2q13del(NPHP1), 15q11.2 dup, 16p11.2 del | 1 | 3 | 1 | 0 | 0 |
| CONGENITAL MALFORMATIONS | 3 | 22q11.2 deletion, 2q13 (NPHP1) deletion, 15q11.2 deletion | 1 | 0 | 0 | 0 | 0 |
| LEARNING DISORDER | 8 | TAR dup; 1q21.1 del; 2q13 (NPHP1) dup, 2q13 (NPHP1) dup, 15q11.2 dup, 15q11.2 dup, 16p11.2 distal del; 22q11.2 del | 8 | 0 | 0 | 0 | 0 |
| HISTORY OF SPECIAL EDUCATION | 4 | 2q13(NPHP1) dup, 15q11.2dup, 1q21.1del, 22q11.2 del | 3 | 10 | 8 | 1 | 1 |
| EXTREME LEARNING DIFFICULTIES (WITHOUT LEARNING DISORDER DIAGNOSIS) | 4 | TAR dup; 15q11.2 del; 16p13.11 del; 22q11.2 dup | 4 | 2 | 2 | 0 | 0 |

Notes: The number of participants affected by each feature, as determined by study assessments, is shown. For NDD CNV carriers, corresponding CNV loci are indicated. One control participant reported a very limited history of special education but without a diagnosed learning disorder. Discordance between EHR data and study-assessed features is noted. del = deletion; dup = duplication.

A previous report detailed medical findings of the Bio*Me* NDD CNV cohort based on EHR diagnostic codes. Current study assessments qualitatively extended those analyses and corroborated medical findings reported in other cohorts of CNV carriers, including obesity, hypertension, neuropathy, and congenital anomalies (Supplementary Table 3). No congenital anomalies were observed in participants with schizophrenia or controls. No participants in this study had short stature.

Regarding functional status, all NDD CNV carriers and controls reported a history of employment without disability, whereas 20% of the schizophrenia group reported disability and were unemployed. All CNV carriers and controls were able to live independently, while 20% of the schizophrenia group lived in supervised residences or were unable to live independently.

### Neurocognitive Assessments

For cognitive analyses, the NDD CNV group was restricted to loci reported in previous UK Biobank analyses to facilitate cross-study comparability, excluding the common 15q13.3 (CHRNA7) duplication, 17p12 deletion, and 2q13 (NPHP1) deletion/duplication, yielding 22 carriers across 11 unique loci.^18, 19^ The 22 NDD CNV carriers were significantly impaired compared to controls on digit span backwards (Beta=-1.76, FDR=0.04) and digit span sequencing (Beta=-2.01, FDR=0.04), but outperformed the schizophrenia group on HVLT-R Immediate Recall (Beta=4.5, FDR=0.05) (Figure 2, Supplementary Table 4). Ancestry was correlated with social cognition (r^2^=0.36, p=6.5x10^-7^), while sex was correlated with social cognition (r^2^=0.12, p=0.003) and verbal learning (r^2^=0.11, p=0.004). Performance of each NDD CNV carrier across seven cognitive domains is summarized, with variation observed by locus (Supplementary Table 5, Supplementary Figure 2). For example, the 22q11.2 deletion carrier was among the lowest performing and the 1q21.1 duplication carrier was among the highest performing.

**Figure 2.**
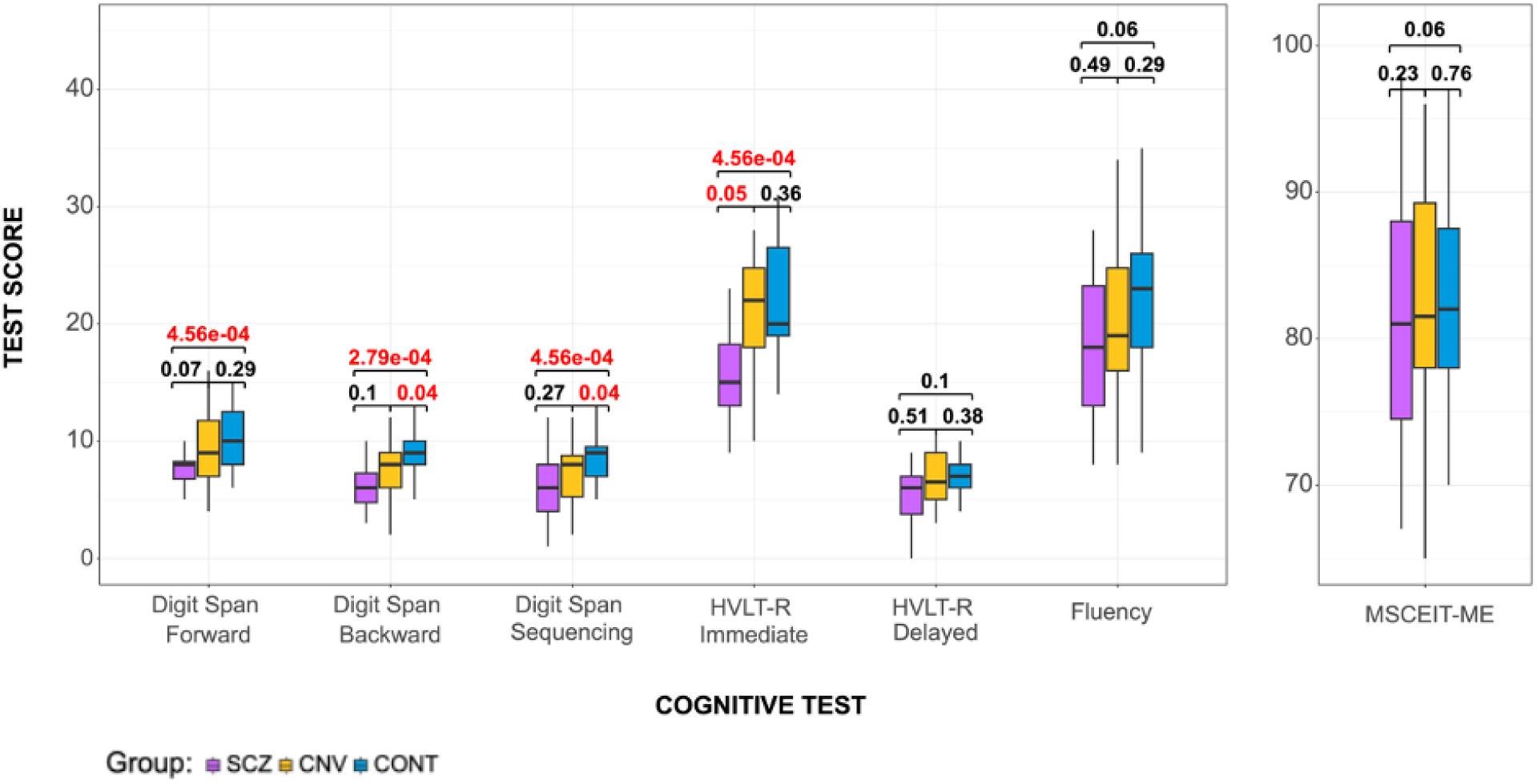
Cognitive Performance by Group. Boxplots showing cognitive test scores across seven cognitive domains for 22 NDD CNV carriers, 20 individuals with schizophrenia, and 23 controls. Each score was modeled by group, adjusting for age, sex, and ancestry. FDR-adjusted p-values are shown; red indicates significance at FDR ≤ 0.05. ***Notes:*** *The x-axis represents cognitive tests, and the y-axis shows raw cognitive test scores. Each score was regressed on group status, adjusting for age, sex, and ancestry. FDR-adjusted values are shown for each test. Red indicates statistical significance at an FDR threshold of 0.05*.

## DISCUSSION

As clinical biobanks expand, opportunities for recall-by-genotype (RbG) will increase across diseases and variant classes, yet practical feasibility remains uncertain given limited published benchmarks. RbG enables direct phenotyping to refine genotype–phenotype relationships and may inform novel, more precise mechanism-based interventions. Notably, within a multi-ancestry healthcare system, this study represents the first RbG investigation of NDD CNV carriers and the first recall of individuals with schizophrenia as comparators. The inclusion of participants from diverse ancestral backgrounds provides insights into RbG participation and generalizability.^24^

In this study, 892 adults were recalled from Bio*Me* for participation. Overall, 18% responded to initial recruitment, 12% were screened, 10% began the study, and 8% completed the study—reflecting challenges in RbG implementation. While the overall recall rate was lower than some previously reported biobank RbG studies, the resulting sample size was comparable to past reports, most of which did not target psychiatric risk variants or include individuals with psychiatric illness (Supplementary Table 6).^33–43^ Factors influencing participation included recent engagement with the healthcare system, participant interest and motivation, and logistical constraints (e.g. scheduling barriers). Recruitment success varied modestly by sex and age, with comparable rates across ancestry and comparator groups, including controls. Other potential influences—such as perceived personal relevance or broader sociocultural factors—were not systematically assessed but warrant evaluation in future studies to inform strategies for improving RbG feasibility and scalability.

Novel findings included the recall of lower-functioning individuals with schizophrenia from the biobank at rates comparable to other groups, including controls. In addition, a marked discordance between study assessments and EHRs highlights the need to validate EHR-derived data in biobank research. In particular, EHRs lacked documentation of developmental histories, whereas study assessments identified developmental findings, retrospectively, among NDD CNV carriers—13% with speech or motor developmental delays, 27% with learning disorders, and 13% with a history of special education. For the NDD CNV carriers, clinical and cognitive findings were broadly consistent with prior, larger studies, most of which comprised predominantly European cohorts, although variable expressivity of NDD CNVs is known.^10, 18, 24, 44, 45^ The current study also assessed social cognition, a domain excluded from some past population cohort studies. Genetic results were not disclosed to participants, consistent with the IRB-approved, research-only design. The initial genotyping was not performed in a CLIA-certified laboratory, precluding a clinical return of results. This approach contrasts with the neuropsychiatric CNV recall report from Geisinger *MyCode* which returned results for CNVs at nine loci.^24^ For the current study, consent and recontact procedures emphasized the study’s observational nature and absence of direct clinical feedback, mitigating ethical concerns while enabling deep phenotyping. Future studies may further address ethical, legal and social (ELSI) challenges of biobank RbG, for which there remains a lack of qualitative ethical research and guidance on disclosing genetic variants, including variants that are not American College of Medical Genetics and Genomics (ACMG) secondary findings or immediately clinically actionable.^46^

Past RbG reports in psychiatry have been limited. The previously published recall of neuropsychiatric CNV carriers differed from the present study in that it was conducted within a relatively contained healthcare system, recruited mostly individuals of European ancestry, did not include cognitive assessments, and offered a return of results.^24^ Another RbG study recalled 197 healthy individuals at extremes of low or high schizophrenia polygenic risk score (PRS) from a population cohort, Avon Longitudinal Study of Parents and Children (ALSPAC), for deep phenotyping, including behavioral assessments and structural and functional imaging.^47^ The study identified an effect of schizophrenia PRS on psychotic experiences, blood oxygen level-dependent signal during reward processing in the ventral striatum and a broader reward-related network, but no effect on whole brain measures or brain morphometry. Another RbG study from ALSPAC recalled 24 individuals who harbored a schizophrenia common risk variant in ZNF804A for deep phenotyping and queried the effect of the risk variant on sleep architecture and memory.^48^ Neither of the RbG studies from ALSPAC returned genetic results.

Beyond psychiatry, the few published biobank RbG studies have mostly focused on genetic variants for cardiometabolic traits or cancer (Supplementary Table 6).^49, 50^ The sample sizes of previous biobank RbG reports have varied by biobank, genomic variant frequency, target population, recall method and study goals, with some studies extending recruitment to family cascade screening.^33–43^ The recall rate of 8% for the current study is lower than some past biobank RbG reports, underscoring the challenges of RbG studies, however a fair number did not report recall rates, precluding direct comparison. Most past RbG studies were descriptive, deploying phenotyping methods for recalled participants, some including remote assessments. One RbG study incorporated Mendelian Randomization analysis to enhance causal inference.^37^ A recent review of biobank RbG studies emphasized that additional RbG studies are “urgently needed” to optimize and leverage the study design, especially for follow-up longitudinal research.^42, 49, 50^

This study had several limitations. The remote administration of assessments, though validated, could have influenced performance, however, it also informs RbG studies for scalability and inclusion of geographically distant participants. The clinical assessments were not specifically designed to evaluate NDD CNV pathogenicity, which has been reported. While some cognitive differences between groups were significant, the modest sample size may have limited the ability to detect smaller effects. Sex and ancestry were included as covariates in pooled analyses, but stratified analyses by these variables were not performed. Some clinical assessments relied on participant self-report and may be subject to recall bias. Although the study excluded medical conditions known to contribute to cognitive impairment, other potential confounders, such as medication burden were not systematically controlled. Analyses focused on a subset of recurrent NDD CNV loci and did not incorporate genome-wide CNV burden or other rare non-recurrent CNVs. NDD CNVs were selected based on prior biobank analyses, with loci of variable effect sizes that may have attenuated group-level findings. Low-functioning NDD CNV carriers may have been underrepresented due to reduced likelihood of study participation, potentially biasing results. The female overrepresentation in the NDD CNV group may limit generalizability; future studies should aim for balanced sex representation. Analyses were pooled across CNV loci and included individuals affected and unaffected by neuropsychiatric or developmental phenotypes, precluding assessment of locus-specific effects. Although not a formal feasibility study, this work offers quantitative benchmarks—spanning response rates, participation patterns, and phenotyping results—that can inform future RbG feasibility and scalability studies.

This study establishes a proof-of-concept for recall-by-genotype of a class of psychiatric risk variants from a multi-ancestry, healthcare-system–derived biobank. The ability to assemble genotypically defined cohorts for follow-up observational or interventional research—both cross-sectional and longitudinal—represents a valuable opportunity for psychiatry. Extending RbG to additional clinical biobanks in healthcare systems, and targeting distinct psychiatric genetic risk variants, could offer a powerful approach to translate disease-relevant genetic findings into clinical insights. Ongoing research evaluating the feasibility, scalability, and generalizability of RbG approaches will be essential to refine genotype–phenotype relationships and advance precision psychiatry.

## Data Availability

The de-identified clinical data summarized herein was obtained from biobank participants in a research study, but is not publicly available, as per IRB-approval and guidance. 

## ACKNOWLEDGEMENTS

We thank individuals within the Bio*Me* Biobank for their participation. We thank colleagues from the Institute of Personalized Medicine at the Icahn School of Medicine for facilitating recall of the BioMe biobank participants (Amanda Merkelson, Sheryl Cruz and Alanna Gomez).

## AUTHOR CONTRIBUTIONS

RB conceptualized the study, obtained funding and secured IRB approval for the project. RB reviewed biobank data, and RB and NZ interacted with recalled study participants to collect additional data. AR contributed to study design and oversight. NZ, BM and RB contributed to data analyses. All authors reviewed and approved the final draft for manuscript.

