## Supplementary Methods and Tables for "Recall-by-Genotype of Neurodevelopmental Disorder Copy Number Variants from a Multi-Ancestry, Healthcare-System Biobank"

*Study Recruitment*: Bio*Me* biobank participants identified for recall for the current study were contacted by both mail and email, with up to two attempts for each modality. If no response was received, participants were subsequently contacted by telephone, with multiple attempts per individual at varying days and times. Recontact was paused if a participant directly expressed a lack of interest in the study or if after six unsuccessful attempts to reach the participant by phone, at variable days and timepoints.

*NDD CNVs*: As previously reported, NDD CNV loci identified in Bio*Me* were culled from a published list of 92 CNVs across 47 genomic regions (including reciprocal deletions and duplications) with varying pathogenicity, as previously described.[1-4]

*EHR/Chart Review:* For the current study, a chart review of each enrolled participant was conducted (by one board-certified psychiatrist, RB) to confirm group assignment (schizophrenia or control) and overall medical history. Records were reviewed from January 1, 2005, through November 12, 2020, including inpatient and outpatient clinical encounter notes, procedure notes, and medication prescription records.

*EHR Visit Density:* Visit density was calculated as the total number of completed clinical encounters with a healthcare provider, from January 1, 2005, through November 12, 2020, with inpatient and outpatient encounters each counted as one visit.

*Clinical Assessment:* The clinical assessment consisted of a validated, structured diagnostic interview, the Mini International Neuropsychiatric Interview (MINI 7.0.2), compatible with DSM-5 and ICD-10 criteria.[5] The MINI (v7.0.2) has been used reliably in telephone-based research with strong concordance between remote and face-to-face diagnostic interviews.[6] In addition, an unstructured clinical interview was conducted to obtain a medical and psychiatric history (including medication use and prior hospitalizations), as well as a retrospective developmental history, and social and occupational histories to assess functional status.

*Neurocognitive Assessment:* While some comprehensive cognitive batteries are well-validated, they are time-intensive and include measures less directly aligned with the cognitive domains targeted in this study. To prioritize assessment of cognitive domains most relevant to our study objectives, we selected individual subtests from established instruments (MATRICS, HVLT-R, WAIS-IV), covering attention, executive function, verbal learning and memory, and social cognition. This approach ensured both psychometric rigor and feasibility for a 30-minute assessment, including remote administration. The neurocognitive battery comprised seven subtests: Digit Span (Forward, Backward, and Sequencing) to assess attention and executive function; Category Fluency to assess processing speed and semantic memory; the Mayer-Salovey-Caruso Emotional Intelligence Test – Managing Emotions branch (MSCEIT-ME) to assess social cognition; and the Hopkins Verbal Learning Test – Revised (HVLT-R) to assess immediate and delayed verbal memory.[7, 8] The immediate recall score represented the total number of words recalled across three learning trials, consistent with the MATRICS Consensus Cognitive Battery (MCCB) protocol. The MSCEIT was scored using the standardized MSCEIT software.[9]

*Remote Assessment*: Each of two 30-minute assessments was conducted at two variable, non-contiguous timepoints, and scheduled as per participant preference. Of the 73 participants who completed both study assessments, 79% participated by phone and 21% by Zoom. Most participants opted for phone (60% of NDD CNV carriers, 95% of schizophrenia group and 91% of controls).

*Remote Assessment Justification*: Accumulating evidence supports the feasibility and validity of remote administration of the clinical and cognitive scales used in this study, including telephone-based assessments among individuals with schizophrenia and cognitive impairments: Structured diagnostic interviews, such as the Mini International Neuropsychiatric Interview (MINI v7.0.2), have been successfully administered remotely, yielding reliable diagnostic classifications comparable to in-person assessments.[10, 11] Measures of attention and executive function, including Digit Span Forward, Backward, and Sequencing, demonstrate acceptable reliability and validity when administered via phone or videoconference in psychiatric populations.[12-14] Processing speed and semantic memory, as assessed by Category Fluency, have similarly shown robust reliability between remote and face-to-face administration.[15, 16] Verbal learning and memory, assessed with the Hopkins Verbal Learning Test–Revised (HVLT-R) for immediate and delayed recall, also maintain good reliability and validity in remote contexts.[16] These findings are consistent with published telepractice guidance for cognitive assessment, including WAIS-IV subtests, which emphasize standardized instructions, environmental control, and attentional monitoring to optimize reliability and validity in remote administration.[17]

**Supplementary Figure 1. Study Assessments, Overview.** Overview of clinical and cognitive assessments administered to study participants.

| CLINICAL ASSESSMENT |
|---|
| Mini International Neuropsychiatric Interview (MINI) |
| Medical History |
| Psychiatric History (including medications and hospitalizations) |
| Developmental History |
| Social, Occupational and Educational History |

| COGNITIVE ASSESSMENT | | |
|---|---|---|
| **Test** | **Domain** | **Battery** |
| Digit Span Forward | Attention | WAIS-IV |
| Digit Span Backward | Executive function, Verbal working memory | WAIS-IV |
| Digit Span Sequencing | Working memory maintenance | WAIS-IV |
| Category fluency | Processing speed | MCCB |
| Hopkins Verbal Learning Test - Revised (HVLT-R) | Delayed recall | MCCB |
| Hopkins Verbal Learning Test - Revised (HVLT-R) | Immediate recall | MCCB |
| MSCEIT-ME | Social Cognition, Emotional intelligence | MCCB |

***Notes:*** *MCCB = MATRICS Consensus Cognitive Battery; WAIS = Wechsler Adult Intelligence Scale*

**<u>Supplementary Figure 2. Cognitive Performance by NDD CNV locus</u>**. Plot showing the cognitive performance rank of each NDD CNV carrier across seven cognitive tests

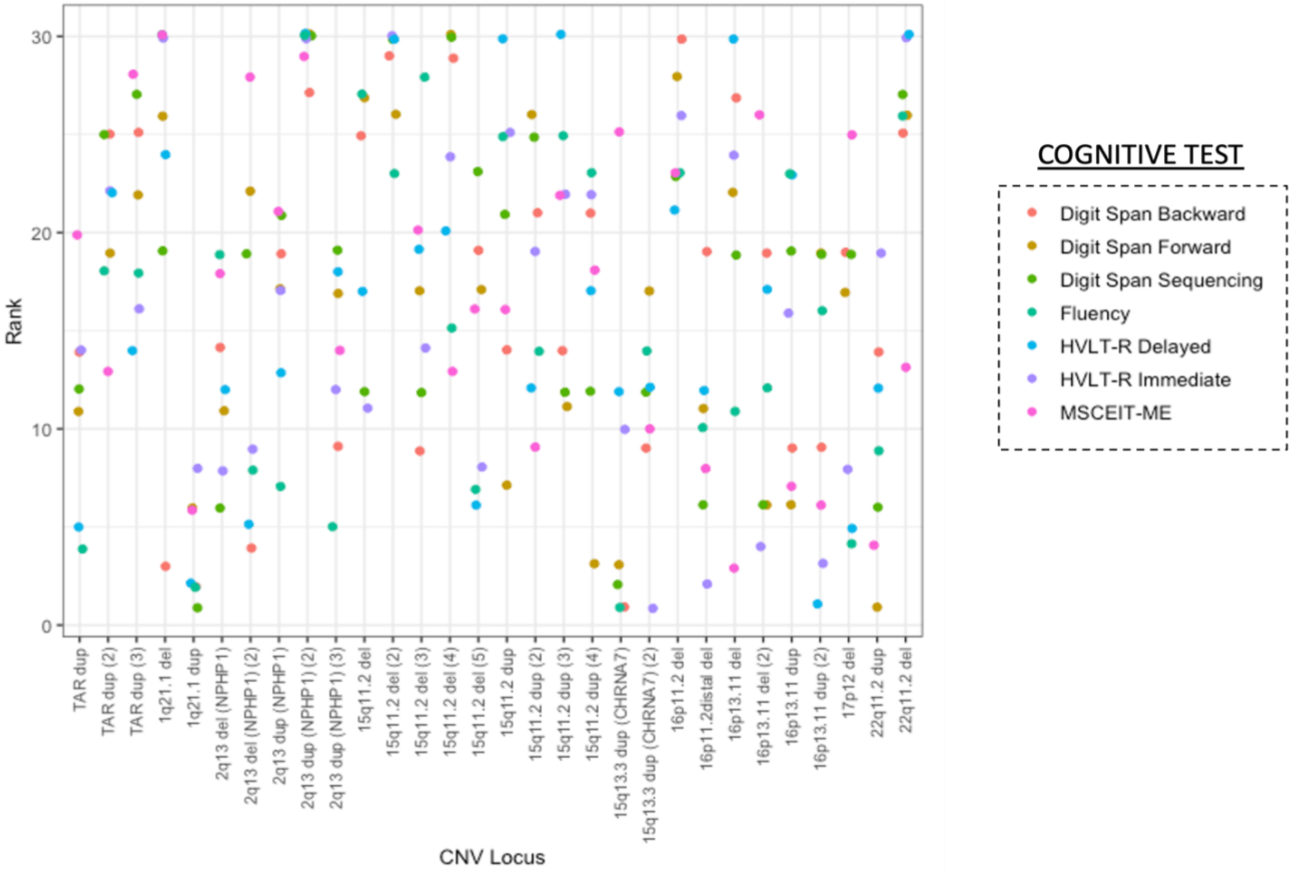


*Notes: See Supplementary Table 4 for ranked loci in tabulated form.*
*This x-axis represents NDD CNV locus. For NDD CNV loci with more than one carrier, sequential numbers are indicated in parentheses, such as (1,2,3). The y-axis reflects rank, where 1=highest performing and 30=lowest (most impaired).*
*Digit Span and HVLT-R were adjusted for age, sex, race/ethnicity, and education. Scores for Fluency and MSCEIT were adjusted for age, sex, and education.*
*del=deletion; dup=duplication.*

**Supplementary Table 1: Study Sample Demographics**. Age, sex and ancestry of biobank participants who were recontacted and completed study.

| | | | RECONTACTED FROM BIOBANK | | | COMPLETED STUDY ASSESSMENTS | | | | COMPLETED vs RECONTACTED, Statistics | | |
|---|---|---|---|---|---|---|---|---|---|---|---|---|
| | Biobank Participants (n=24,877) | NDD CNV (n=627) | NDD CNV (n=335) | Schizophrenia (n=217) | Controls (n=340) | NDD CNV (n=30) | Schizophrenia (n=20) | Controls (n=23) | Total (n=73) | NDD -CNV | SCZ | CONT |
| Mean Age, Biobank Enrollment (SD) | 50.5 (17.3) | 50.4 (17.0) | 39.0 (10.8) | 45.4 (9.9) | 41.9 (10.7) | 37.3 [9.2] | 41 [9.3] | 39.9 [9.4] | | t=0.87, p=0.39 | t=2.03, p=0.05 | t=0.85, p=0.40 |
| Mean Age at Study Assessment (SD) | | | | | | 47.3 [9.5] | 50.2 [10.1] | 49.5 [11.2] | 48.8 [10.2] | | | |
| Female (%) | 14,586 (59%) | 389 (62%) | 219 (65%) | 97 (45%) | 209 (61%) | 26 (87%) | 8 (40%) | 14 (61%) | 48 (66%) | X2=5.9, p=0.02 | X2=0.1, p=0.78 | X2=0.00, p=0.99 |
| Ancestry (%) | | | | | | - | - | - | | X2=1.4, p=0.70 | X2=2.1, p=0.55 | X2=2.8, p=0.43 |
| African | 5965 | 168 | 95 | 101 | 111 | 10 (33%) | 9 (45%) | 8 (35%) | 27 (37%) | | | |
| European | 7892 | 208 | 102 | 17 | 52 | 10 (33%) | 2 (10%) | 7 (30%) | 19 (26%) | | | |
| Hispanic | 8536 | 200 | 109 | 87 | 151 | 9 (30%) | 9 (45%) | 7 (30%) | 25 (34%) | | | |
| Other | 2484 | 51 | 32 | 12 | 26 | 1 (3%) | 0 (0) | 1 (4%) | 2 (3%) | | | |

*Notes: p values reflect comparisons between participants and nonparticipants within each group; Chi-square values for sex and ancestry are indicated, and t-test is for age. Ancestry is as per self-reported*

**Supplementary Table 2: NDD CNV Loci, Prevalence**. Counts of NDD CNV carriers, by locus, with chromosomal coordinates indicated.

| NDD CNV | hg19 coordinates | Genes (n) | BioMe (n=627) | Recontacted (n=335) | Completed Study (n=30) | NDD CNV Prevalence: BioMe Total vs Study (p-value) |
|---|---|---|---|---|---|---|
| TAR_del | chr1:145,394,955-145,807,817 | 17 | 5 | 2 | | |
| TAR_dup | chr1:145,394,955-145,807,817 | 17 | 18 | 8 | 3 | 2.29E-05 |
| 1q21.1del** | chr1:146,527,987-147,394,444 | 9 | 8 | 5 | 1 | 0.27 |
| 1q21.1dup | chr1:146,527,987-147,394,444 | 9 | 3 | 2 | 1 | 0.05 |
| NRXN1_DEL** | chr2:50145643-51259674 | 1 | 3 | 1 | | |
| 2q11.2del | chr2:96,742,409-97,677,516 | 22 | 2 | 1 | | |
| 2q11.2dup | chr2:96,742,409-97,677,516 | 22 | 1 | 1 | | |
| 2q13del(NPHP1) | chr2:110,862,716-110,983,948 | 3 | 81 | 38 | 2 | 0.70 |
| 2q13dup(NPHP1) | chr2:110,862,716-110,983,948 | 3 | 62 | 32 | 3 | 0.08 |
| 2q13del | chr2:111,394,040-112,012,649 | 3 | 3 | 3 | | |
| 2q13dup | chr2:111,394,040-112,012,649 | 3 | 8 | 7 | | |
| 2q21.1del | chr2:131,481,308-131,930,677 | 5 | 9 | 6 | | |
| 2q21.1dup | chr2:131,481,308-131,930,677 | 5 | 5 | 3 | | |
| 3q29del** | chr3:195,720,167-197,354,826 | 28 | 1 | 1 | | |
| 3q29dup | chr3:195,720,167-197,354,826 | 28 | 0 | 0 | | |
| Sotos_5q35del | chr5:175,720,924-177,052,594 | 39 | 0 | 0 | | |
| 5q35dup | chr5:175,720,924-177,052,594 | 39 | 0 | 0 | | |
| 7q11.23_del | chr7:72,744,915-74,142,892 | 26 | 1 | 0 | | |
| 7q11.23_dup | chr7:72,744,915-74,142,892 | 26 | 0 | 0 | | |
| 7q11.23dup_distal | chr7:75,138,294-76,064,412 | 16 | 0 | 0 | | |
| 8p23.1del | chr8:8,098,990-11,872,558 | 35 | 0 | 0 | | |
| 8p23.1dup | chr8:8,098,990-11,872,558 | 35 | 0 | 0 | | |
| 10q11.21q11.23del | chr10:49,390,199-51,058,796 | 19 | 1 | 0 | | |
| 10q11.21q11.23dup | chr10:49,390,199-51,058,796 | 19 | 2 | 0 | | |
| 10q23del | chr10:82,045,472-88,931,651 | 29 | 0 | 0 | | |
| 10q23dup | chr10:82,045,472-88,931,651 | 29 | 1 | 0 | | |
| 13q12del(CRYL1) | chr13:20,977,806-21,100,012 | 2 | 8 | 4 | | |
| 13q12dup(CRYL1) | chr13:20,977,806-21,100,012 | 2 | 0 | 0 | | |
| 13q12.12del | chr13:23,555,358-24,884,622 | 10 | 4 | 3 | | |
| 13q12.12dup | chr13:23,555,358-24,884,622 | 10 | 9 | 3 | | |
| 15q11.2del | chr15:22,805,313-23,094,530 | 5 | 60 | 27 | 5 | 8.01E-05 |
| 15q11.2dup | chr15:22,805,313-23,094,530 | 5 | 163 | 89 | 4 | 0.39 |
| PW/AS_15q11.2q13.1 BP1-3_del | chr15:22,805,313-28390339 | 116 | 0 | 0 | | |
| PW/AS_15q11.2q13.1 BP1-3_dup | chr15:22,805,313-28390339 | 116 | 0 | 0 | | |
| 15q11q13del_BP3-BP4(APBA2_TJP) | chr15:29,161,368-30,375,967 | 4 | 1 | 1 | | |
| 15q11q13dup_BP3-BP4(APBA2_TJP) | chr15:29,161,368-30,375,967 | 4 | 3 | 1 | | |
| 15q11q13dup_BP3-BP5 | chr15:29,161,368-32462776 | 17 | 0 | 0 | | |
| 15q13.3del | chr15:31,080,645-32,462,776 | 8 | 5 | 0 | | |
| 15q13.3dup | chr15:31,080,645-32,462,776 | 8 | 5 | 2 | | |
| 15q13.3del(CHRNA7) | chr15:32,017,070-32,453,068 | 1 | 5 | 2 | | |
| 15q13.3dup(CHRNA7) | chr15:32,017,070-32,453,068 | 1 | 42 | 19 | 2 | 0.23 |
| 15q24del | chr15:72900171-78151253 | 77 | 0 | 0 | | |
| 15q24dup | chr15:72900171-78151253 | 77 | 0 | 0 | | |
| 16p13.11del | chr16:15,511,655-16,293,689 | 7 | 13 | 4 | 2 | 3.35E-03 |
| 16p13.11dup | chr16:15,511,655-16,293,689 | 7 | 39 | 19 | 2 | 0.19 |
| 16p12.1del | chr16:21,950,135-22,431,889 | 8 | 2 | 0 | | |
| 16p12.1dup | chr16:21,950,135-22,431,889 | 8 | 14 | 7 | | |
| 16p11.2distal_del** | chr16:28,823,196-29,046,783 | 11 | 5 | 4 | 1 | 0.13 |
| 16p11.2distal_dup | chr16:28,823,196-29,046,783 | 11 | 4 | 3 | | |
| 16p11.2del | chr16:29,650,840-30,200,773 | 30 | 15 | 12 | 1 | 0.53 |
| 16p11.2dup** | chr16:29,650,840-30,200,773 | 30 | 4 | 4 | | |
| 17p12del | chr17:14,141,387-15,426,961 | 8 | 6 | 5 | 1 | 0.18 |
| 17p12dup | chr17:14,141,387-15,426,961 | 8 | 10 | 7 | | |
| Smith Magenis Syndrome_17p11.2del | chr17:16,812,771-20,211,017 | 59 | 0 | 0 | | |
| Potocki-Lupski syndrome_17p11.2dup | chr17:16,812,771-20,211,017 | 59 | 0 | 0 | | |
| 17q11.2del(NF1) | chr17:29,107,491-30,265,075 | 19 | 0 | 0 | | |
| 17q11.2dup(NF1) | chr17:29,107,491-30,265,075 | 19 | 0 | 0 | | |
| 17q12del | chr17:34,815,904-36,217,432 | 17 | 4 | 2 | | |
| 17q12dup | chr17:34,815,904-36,217,432 | 17 | 4 | 2 | | |
| 17q21.31del | chr17:43,705,356-44,164,691 | 10 | 0 | 0 | | |
| 22q11.2del** | chr22:19,037,332-21,466,726 | 61 | 1 | 1 | 1 | 3.46E-03 |
| 22q11.2dup | chr22:19,037,332-21,466,726 | 61 | 7 | 4 | 1 | 0.22 |
| 22q11.2distal_del | chr22:21,920,127-23,653,646 | 26 | 0 | 0 | | |
| 22q11.2distal_dup | chr22:21,920,127-23,653,646 | 26 | 0 | 0 | | |

***=NDD CNV loci previously reported to be significantly associated with schizophrenia by GWAS (Marshall CR, et al. Contribution of copy number variants to schizophrenia from a genome-wide study of 41,321 subjects. Nat Genet. 2017;49(1):27-35).*

*p-value is for a Test of Equal Proportions*

**Supplementary Table 3: Selected Medical Features, by NDD CNV locus**. Overlap of medical features identified during current assessment with conditions previously reported in other larger-scale cohorts.

| CNV locus | Location (hg19) | BioMe CNV Sample (n) | Medical Phenotypes overlapping UK Biobank or Previous Reports* |
|---|---|---|---|
| TAR_dup | chr1:145,39-145,81 | 3 | Obesity |
| 1q21.1del | chr1:146,53-147,39 | 1 | |
| 1q21.1dup | chr1:146,53-147,39 | 1 | Gastric Ulcers |
| 2q13del (*NPHP1* ) | chr2:110,86-110,98 | 2 | Congenital Cardiac Anomaly |
| 2q13dup (*NPHP1* ) | chr2:110,86-110,98 | 3 | |
| 15q11.2del | chr15:22,81-23,09 | 5 | Congenital renal anomaly |
| 15q11.2dup | chr15:22,81-23,09 | 4 | |
| 15q13.3dup (*CHRNA7* ) | chr15:32,02-32,45 | 2 | |
| 16p13.11del | chr16:15,51-16,29 | 2 | |
| 16p13.11dup | chr16:15,51-16,29 | 2 | Hypertension |
| 16p11.2distal_del | chr16:28,82-29,05 | 1 | Obesity |
| 16p11.2del | chr16:29,65-30,20 | 1 | Obesity, Hypertension |
| 17p12del(HNPP) | chr17:14,14-15,43 | 1 | Neuropathy |
| 22q11.2del | chr22:19,04-21,47 | 1 | Congenital Cardiac Anomaly, Thrombytopenia |
| 22q11.2dup | chr22:19,04-21,47 | 1 | |

**Complete medical histories of NDD CNV carriers are not indicated, but rather only conditions that overlap previous reports in other cohorts, as summarized in Crawford K. et al. Medical consequences of pathogenic CNVs in adults: analysis of the UK Biobank. J Med Genet. 2019;56(3):131-8 Medical phenotypes during current study were determined by a consensus of chart review, clinical interview and patient self-report*

*For statistical associations of NDD CNVs with ICD codes, see Birnbaum et al. Clinical Characterization of Copy Number Variants Associated With Neurodevelopmental Disorders in a Large-scale Multiancestry Biobank. JAMA Psychiatry. 2022;79(3):250-9.*

**Supplementary Table 4: Cognitive Effects of NDD CNVs**: Results of linear regression analyses of seven cognitive tests, compared by group status.

**NDD CNV subset (n=22) compared to schizophrenia (n=20) and controls (n=23)**

| Cogntive Test | β | | t-value | | p-value | | FDR | |
|---|---|---|---|---|---|---|---|---|
| | Controls | Schizophrenia | Controls | Schizophrenia | Controls | Schizophrenia | Controls | Schizophrenia |
| DigitSpan_Forward | 1.10 | -2.00 | 1.41 | -2.38 | 0.16 | 0.02 | 0.29 | 0.07 |
| DigitSpan_Backward | 1.76 | -1.54 | 2.57 | -2.08 | 0.01 | 0.04 | **0.04** | 0.10 |
| DigitSpan_Sequencing | 2.01 | -1.10 | 2.60 | -1.33 | 0.01 | 0.19 | **0.04** | 0.27 |
| HVLTR_Immediate | 1.69 | -4.50 | 1.14 | -2.79 | 0.26 | 0.01 | 0.36 | **0.05** |
| HVLTR_Delayed | 0.80 | -0.57 | 0.99 | -0.66 | 0.33 | 0.51 | 0.38 | 0.51 |
| Fluency | 2.82 | -1.73 | 1.44 | -0.82 | 0.16 | 0.42 | 0.29 | 0.49 |
| MSCEIT-ME | 0.63 | -3.37 | 0.31 | -1.53 | 0.76 | 0.13 | 0.76 | 0.23 |

*Reference group: CNV*

**Schizophrenia (n=20) vs Controls (n=23)**

| Cogntive Test | β | t-value | p-value | FDR |
|---|---|---|---|---|
| DigitSpan_Forward | -3.10 | -3.89 | 2.30E-04 | **4.56E-04** |
| DigitSpan_Backward | -3.24 | -4.40 | 3.99E-05 | **2.79E-04** |
| DigitSpan_Sequencing | -3.07 | -3.86 | 2.60E-04 | **4.56E-04** |
| HVLTR_Immediate | -6.12 | -3.93 | 2.02E-04 | **4.56E-04** |
| HVLTR_Delayed | -1.33 | -1.65 | 0.10 | 0.10 |
| Fluency | -4.39 | -2.07 | 0.04 | 0.06 |
| MSCEIT-ME | -4.03 | -1.97 | 0.05 | 0.06 |

***Reference group: Controls***

*Notes: β coefficients from linear models adjusted for age, sex, and ancestry. p values are FDR-corrected for multiple comparisons (FDR ≤ 0.05 shown in bold).*

**Supplementary Table 5. Cognitive Test Performance Rank, by NDD CNV Locus**. Rank of cognitive scores by NDD CNV locus.

| NDD CNV Locus | Overall_Rank | Digit Span Forward | Digit Span Backward | Digit Span Sequencing | HVLT-R Immediate | HVLT-R Delayed | Fluency | MSCEIT-ME |
|---|---|---|---|---|---|---|---|---|
| 1q21.1 dup | 1 | 6 | 2 | 1 | 8 | 2 | 2 | 6 |
| 15q13.3 dup (CHRNA7) | 2 | 3 | 1 | 2 | 10 | 12 | 1 | 25 |
| 22q11.2 dup | 3 | 1 | 14 | 6 | 19 | 12 | 9 | 4 |
| 16p11.2distal del | 4 | 11 | 19 | 6 | 2 | 12 | 10 | 8 |
| 16p13.11 dup (2) | 5 | 19 | 9 | 19 | 3 | 1 | 16 | 6 |
| 15q13.3 dup (CHRNA7) (2) | 6 | 17 | 9 | 12 | 1 | 12 | 14 | 10 |
| TAR dup | 7 | 11 | 14 | 12 | 14 | 5 | 4 | 20 |
| 2q13 del (NPHP1) | 8 | 11 | 14 | 6 | 8 | 12 | 19 | 18 |
| 16p13.11 del (2) | 9 | 6 | 19 | 6 | 4 | 17 | 12 | 26 |
| 2q13 dup (NPHP1) (3) | 10 | 17 | 9 | 19 | 12 | 18 | 5 | 14 |
| 2q13 del (NPHP1) (2) | 11 | 22 | 4 | 19 | 9 | 5 | 8 | 28 |
| 15q11.2 del (5) | 12 | 17 | 19 | 23 | 8 | 6 | 7 | 16 |
| 17p12 del | 13 | 17 | 19 | 19 | 8 | 5 | 4 | 25 |
| 16p13.11 dup | 14 | 6 | 9 | 19 | 16 | 23 | 23 | 7 |
| 2q13 dup (NPHP1) | 15 | 17 | 19 | 21 | 17 | 13 | 7 | 21 |
| 15q11.2 dup (4) | 16 | 3 | 21 | 12 | 22 | 17 | 23 | 18 |
| 15q11.2 del (3) | 17 | 17 | 9 | 12 | 14 | 19 | 28 | 20 |
| 15q11.2 dup (2) | 18 | 26 | 21 | 25 | 19 | 12 | 14 | 9 |
| 15q11.2 dup (3) | 20 | 11 | 14 | 12 | 22 | 30 | 25 | 22 |
| 16p13.11 del | 20 | 22 | 27 | 19 | 24 | 30 | 11 | 3 |
| 15q11.2 dup | 21 | 7 | 14 | 21 | 25 | 30 | 25 | 16 |
| 15q11.2 del | 22 | 27 | 25 | 12 | 11 | 17 | 27 | NA |
| TAR dup (2) | 23 | 19 | 25 | 25 | 22 | 22 | 18 | 13 |
| TAR dup (3) | 24 | 22 | 25 | 27 | 16 | 14 | 18 | 28 |
| 15q11.2 del (4) | 25 | 30 | 29 | 30 | 24 | 20 | 15 | 13 |
| 1q21.1 del | 26 | 26 | 3 | 19 | 30 | 24 | 30 | 30 |
| 16p11.2 del | 27 | 28 | 30 | 23 | 26 | 21 | 23 | 23 |
| 22q11.2 del | 28 | 26 | 25 | 27 | 30 | 30 | 26 | 13 |
| 15q11.2 del (2) | 29 | 26 | 29 | 30 | 30 | 30 | 23 | NA |
| 2q13 dup (NPHP1) (2) | 30 | 30 | 27 | 30 | 30 | 30 | 30 | 29 |

*Note: For ranking of Digit Span tests and HVLT-R, cognitive scores were adjusted for age, sex, race/ancestry and years of education . For ranking of Fluency and MSCEIT-ME scores were adjusted for age, sex, and years of education. (See Supplementary Methods)*

*A rank of '1' is highest performing and '30' is lowest (most impaired). See Supplementary Figure 1 for a plot of rankings, by NDD CNV carrier and by cognitive test*

*Parentheses indicate the number of carriers if more than one carrier*

**<u>Supplementary Table 6. Recall-by-Genotype Studies in Biobanks (Non-Psychiatric).</u>** Summary of RbG of non-psychiatric risk variants, showing target variant, recall rates, sample sizes, and main findings.

| Disease Area | Genetic Variant(s) | Biobank | Study Design | Number of Participants | Recall Rate | Main Results | Genetic Disclosure / Return-of-Results? | PubMed ID |
|---|---|---|---|---|---|---|---|---|
| Diabetes | IL2RA haplotypes (rs12722495, rs11594656, rs2104286) | Cambridge BioResource | Recalled individuals by haplotype to measure IL2RA surface expression on T cells; flow cytometry | ~200 per haplotype group | NR | Protective haplotype had increased IL2RA expression on memory CD4+ T cells; functional link to T1D risk | No | 19701192 |
| Breast, Ovarian Cancer | BRCA1 / BRCA2 pathogenic variants | Estonian Biobank | RbG for clinical follow-up, genetic counseling, | 40 | 55% | 22 responded and received results; 8 high-risk, 5 underwent risk-reducing surgery, 10 adhered to surveillance, 10 relatives screened | Yes | 33230308 |
| Familial Hypercholesterolemia (FH) | Rare variants pathogenic for FH in LDLR / APOB / PCSK9 | Estonian Biobank | RbG for clinical/lipid testing + genetic counseling | 27 probands + 20 family members (Cascade screening) | 70% | Reclassified 51% of the study participants from nonspecific hypercholesterolemia to having FH; 32% were unaware of harboring a high-risk disease-associated genetic variant | Yes | 35928446 |
| Wilson's Disease | ATP7B variants | Estonian Biobank | RbG for deep phenotyping (clinical assessment) and serum copper measurements | 17 | 88% | Identified underdiagnosed WD carriers, many with early neurodegenerative signs | Yes | 39674827 |
| Body Mass Index and Cardiovascular Health | BMI GRS extremes | ALSPAC | RbG at Age 21 for top vs bottom BMI GRS percentiles for cardiovascular phenotyping | 418 proposed | 20% | Differences in BMI, SBP across extremes; genotype-based stratification reduced confounding | No | 30524135 |
| Smoking / Nicotine Metabolism | CHRNA5-A3-B4 genotype | ALSPAC | RbG to compare cotinine levels, smoking topography | 200 proposed | NR | Test genotype-phenotype relationships in smoking | No | 24451018 |
| Adiponectin / Metabolic Traits | ADIPOQ variants | Exeter 10,000 | RbG of carriers vs non-carriers for metabolic phenotyping | 12 | NR | Functional genomic investigations of adiponectin biology (i.e. ADIPOQ splicing) | No | 26996131 |
| Obesity | AMY1 copy number variation | DESIR | RbG to compare metabolomic signatures of high- and low-copy number carriers, Measured serum pancreatic and total amylase levels | 100 | NR | Identified differences in lipid metabolism between the two groups | No | 35928446 |
| Alzheimer's Disease | Alzheimer's Disease (AD) risk SNPs | Finnish National Biobank | RbG for digital cognitive and remote assessments | 27 | 19% | Identified cognitive deficits and blood-based biomarkers distinguishing AD from mild cognitive disorder. | No | 37537264 |
| Alzheimer's Disease | AD-Polygenic Risk Score extremes | PROTECT | RbG of individuals at extremes of Alzheimer's PRS for memory and imaging assessments | 16 | NR | Associations with memory deficits and cingulate cortex differences | No | 38087383 |

*Notes: RbG = recall-by-genotype; NR=Not reported*
